# TRACKing Excess Deaths (TRACKED) – an interactive online tool to monitor excess deaths associated with COVID-19 pandemic in the United Kingdom

**DOI:** 10.1101/2020.06.05.20121962

**Authors:** Michael TC Poon, Paul M Brennan, Kai Jin, Jonine D Figueroa, Cathie LM Sudlow

**Author notes:** **Correspondence to:** Professor Cathie LM Sudlow.

## Abstract

**Background:** We aimed to describe trends of excess mortality in the United Kingdom (UK) stratified by nation and cause of death, and to develop an online tool for reporting the most up to date data on excess mortality.

**Methods:** Population statistics agencies in the UK including the Office for National Statistics (ONS), National Records of Scotland (NRS), and Northern Ireland Statistics and Research Agency (NISRA) publish weekly data on deaths. We used mortality data up to 22^nd^ May in the ONS and the NISRA and 24^th^ May in the NRS. Crude mortality for non-COVID deaths (where there is no mention of COVID-19 on the death certificate) calculated. Excess mortality defined as difference between observed mortality and expected average of mortality from previous 5 years.

**Results:** There were 56,961 excess deaths and 8,986 were non-COVID excess deaths. England had the highest number of excess deaths per 100,000 population (85) and Northern Ireland the lowest (34). Non-COVID mortality increased from 23rd March and returned to the 5-year average on 10th May. In Scotland, where underlying cause mortality data besides COVID-related deaths was available, the percentage excess over the 8-week period when COVID-related mortality peaked was: dementia 49%, other causes 21%, circulatory diseases 10%, and cancer 5%. We developed an online tool (TRACKing Excess Deaths - TRACKED) to allow dynamic exploration and visualisation of the latest mortality trends (http://trackingexcessdeaths.com).

**Conclusions:** Continuous monitoring of excess mortality trends and further integration of age- and gender-stratified and underlying cause of death data beyond COVID-19 will allow dynamic assessment of the impacts of indirect and direct mortality of the COVID-19 pandemic.

## Introduction

Suppression and mitigation strategies adopted by the UK government and public health agencies have been necessary for COVID-19 disease control. In the United Kingdom (UK), 47,975 people have died of COVID-19 in the latest data released in late-May from UK death registries.1–3 However, the impact of these strategies has extended beyond only patients with COVID-19. Patients with non-COVID-19 diagnoses may have been harmed as a result of indirect effects of COVID-19, including changes in health service provision, health behaviours, or socio-economic effects.

Excess mortality is the number of deaths above the expected average from previous years. Analysis of the total excess mortality provides a measure of both direct and indirect effects from COVID-19. In the UK, COVID-19 related deaths (where COVID-19 was mentioned on the death certificate, whether or not it was the primary underlying cause) are a measure of the direct impact of COVID-19.1,2,4 Trends in non COVID-related deaths can demonstrate the possible indirect effects of COVID-19. Excess deaths due to changes in behaviour or access to health services may take longer to accrue depending on the natural history of different diseases. Tracking excess mortality and understanding its determinants will allow development of health policies and services that minimise indirect causes of excess deaths.

Population statistics agencies in the UK publish weekly overall and COVID-19 mortality data.1–3 In Scotland, timely reporting of cause-specific mortality data allows trend assessment of excess deaths stratified by broad underlying cause.5 As the UK begins to ease from full to partial suppression strategy, monitoring of excess deaths across the country will be a crucial component of understanding the impact of measures to control the pandemic. We aimed to describe trends in excess mortality in the UK, stratified by nation and cause of death, and to develop an online tool for reporting the most up to date data on excess mortality. The SARS-CoV-2 pandemic has had significant impact beyond the UK on global health and the approach we describe will be of value worldwide.

## Methods

### Data sources

The Office for National Statistics (ONS), National Records of Scotland (NRS), and Northern Ireland Statistics and Research Agency (NISRA) publish weekly mortality data on Tuesday, Wednesday, and Friday, respectively. The average total deaths of the corresponding week from the previous 5 years are provided and are considered as the expected number of deaths. ONS reports annual population sizes in each of the four nations in the UK. We use these publicly available data for our analyses.

### Definitions

COVID-related deaths refer to deaths where COVID-19 was mentioned on the death certificate.1–3 Non-COVID deaths are deaths without mention of COVID-19 on the death certificate. NRS provides cause-specific deaths categorised using the 10th revision of the International Classification of Disease (ICD-10) codes. These include cancer (C00-C97), dementia/Alzheimer’s (F01, F03, and G30), circulatory diseases including heart disease and stroke (I00-I99), non-COVID respiratory diseases (J00-J99), and others. We define excess deaths as the difference from expected number of deaths.

ONS, NRS, and NISRA have different definitions of week number. The end date of the first week is: 3rd Jan in ONS, 5th Jan in NRS, and 10th Jan in NISRA. The date of the first confirmed COVID-19 case was 31st Jan 2020 in England, 27th Feb 2020 in Northern Ireland, 28th Feb 2020 in Wales, and 1st March 2020 in Scotland. England, Scotland, and Wales adopted full suppression strategy “lockdown” on 26th March 2020 and Northern Ireland followed on 28th March 2020.

### Data processing and statistical procedures

All code is available to view and download at https://github.com/michaelpoontc/tracked. Briefly, the script automatically downloads and extracts the latest data from ONS, NRS, and NISRA. Data is organised into weekly overall deaths, COVID-related deaths, non-COVID deaths, and 5-year average deaths stratified by UK nation. For NRS, data on cause-specific deaths are also extracted.

We calculated the crude number of deaths per 100,000 persons and derived 95% confidence intervals (CI) using the normal approximation to the binomial distribution. To visualise trends, we also calculated 4-week rolling means and CIs. We performed all programming and statistical procedures in R (version 4.0.0). The R packages used are available from the source code.

## Results

These results are based on the latest data release up to 22^nd^ May 2020 in England, Wales, and Northern Ireland and 24^th^ May 2020 in Scotland. The total number of deaths was 323,057, of which 56,961 were excess deaths compared with the 5-year average. There were 47,975 COVID-related deaths, representing the majority (84%) of excess deaths, and 8,986 non-COVID excess deaths. The numbers of deaths per 100,000 population in each nation are presented in Table 1. England had the highest number of all, COVID-related, and non-COVID excess deaths (89, 74 and 15 per 100,000 respectively). Northern Ireland had the lowest number of excess deaths, all COVID-related (34 per 100,000). COVID-related deaths were also the sole contributor to excess deaths in Wales.

**Table 1.** Number of deaths per 100,000 stratified by nations of the United Kingdom from the start of 2020 to late-May^†^.

| Nations | Number of deaths per 100,000 people* |  |  |  |
| --- | --- | --- | --- | --- |
|  | Total<br>N (95% CI) | Excess<br>N (95% CI) | COVID-related excess<br>N (95% CI) | Non-COVID excess<br>N (95% CI) |
| UK | 484 (482-485) | 85 (85-86) | 72 (71-72) | 13 (13-14) |
| England | 480 (478-482) | 89 (89-90) | 74 (74-75) | 15 (15-16) |
| Scotland | 532 (526-538) | 75 (73-78) | 65 (63-68) | 10 (9-11) |
| Wales | 529 (521-537) | 60 (57-63) | 66 (63-69) | -6 (-5 to -7) |
| Northern Ireland | 380 (371-389) | 34 (31-37) | 37 (34-40) | -3 (-2 to -4) |

<sup>†</sup>Data from the ONS for England and Wales covers 28<sup>th</sup> Dec 2019 to 22<sup>nd</sup> May 2020; data from NRS for Scotland covers 30<sup>th</sup> Dec 2019 to 24<sup>th</sup> May 2020; data from NISRA for Northern Ireland covers 3<sup>rd</sup> Jan 2020 to 22<sup>nd</sup> May 2020. \*Confidence intervals calculated using normal approximation to the binomial distribution. COVID-related deaths are deaths where COVID-19 was mentioned on the death certificate. Non-COVID deaths are deaths where there was no mention of COVID-19 on the death certificate.

### Overall mortality trends

In all four nations, mortality was lower than the 5-year average from mid-January to the beginning of March (Figure 1). Mortality rose sharply from the week beginning 23rd March when all UK nations adopted full suppression strategy. Peak mortality occurred between 13th April and 26th April. The overall mortality trends were parallel to the COVID mortality trends.

**Figure 1.**
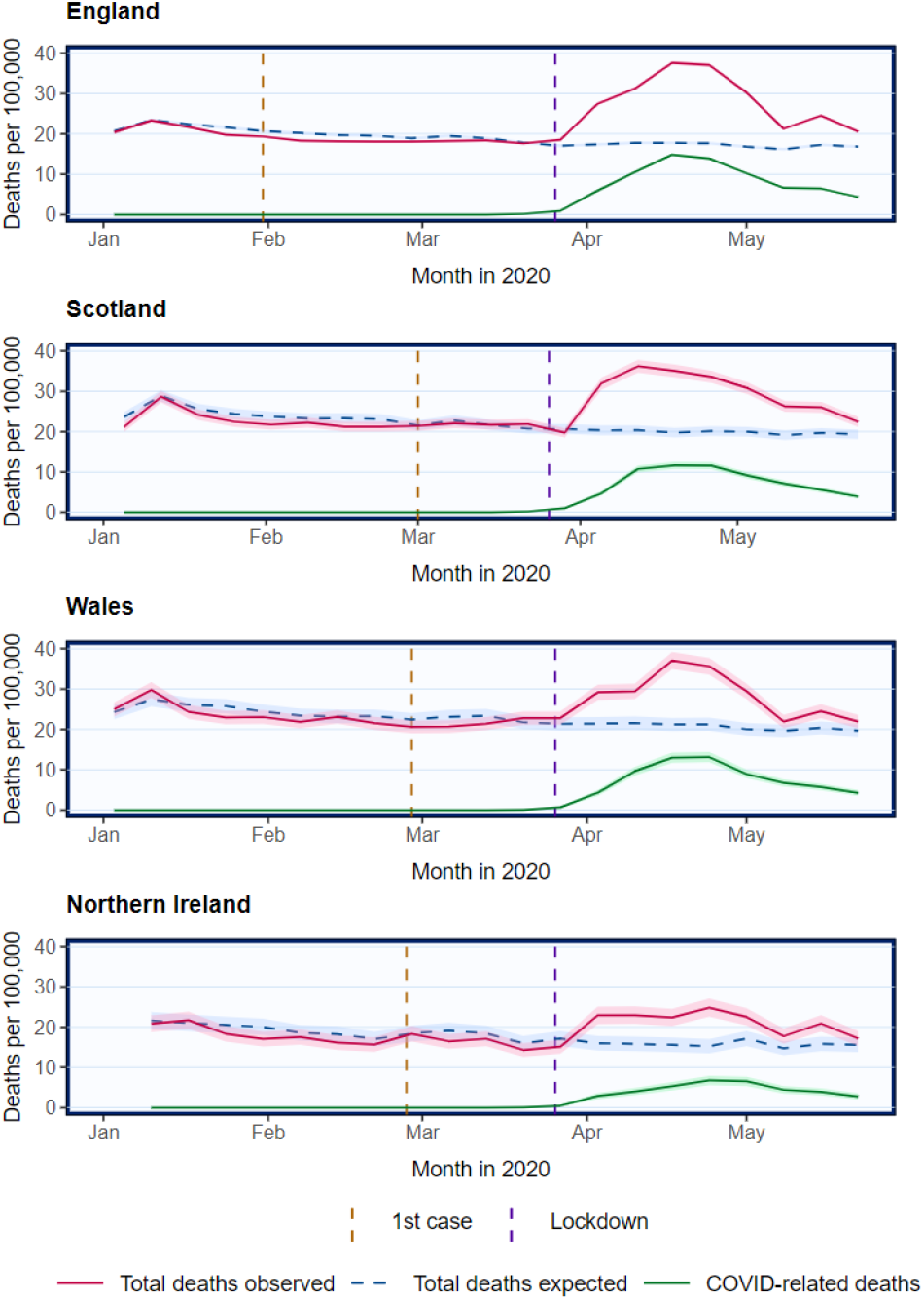
Overall crude mortality trends in the UK from January 2020

### Non-COVID mortality trends

In England and Scotland, non-COVID mortality increased from 23rd March and returned to the 5-year average on 10th May, following the trends of COVID-related deaths (Figure 2). In Wales and Northern Ireland, there was little apparent change in non-COVID mortality with overlapping 95% CI, which may be due to the lower number of deaths reported (Figure 2).

**Figure 2.**
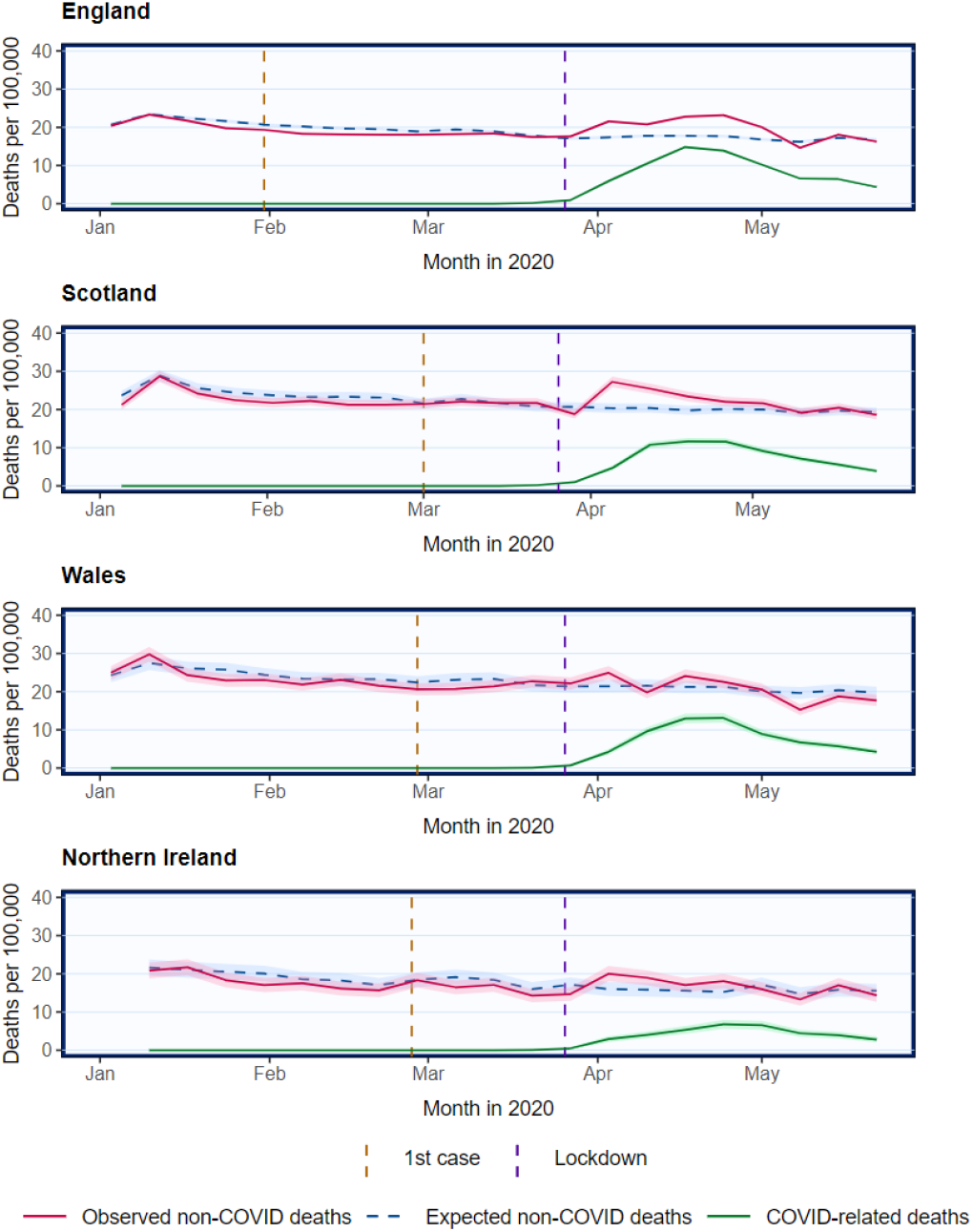
Crude mortality trends of non-COVID and COVID-related deaths in the UK from January 2020

### Cause-specific mortality in Scotland

The latest data from Scotland includes up to 24th May 2020. Overall, in 2020, there were 17% (N = 4,129) excess deaths compared to expected, and 2% (N = 553) excess non-COVID deaths. The number of excess deaths by cause is presented in Table 2. Deaths from dementia and other causes were 15% and 12% higher than the respective number of expected deaths. There were 3% excess deaths attributable to cancer. There was little overall mortality change to circulatory disease. There was a deficit of 20% in non-COVID respiratory deaths.

**Table 2.** Number of deaths in Scotland stratified by cause of death in different periods of time.

| Cause of death† | All available data<br>(30 Dec 2020 to 17 May 2020) |  |  |  | 8-week period over the peak of mortality<br>(23 Mar 2020 to 17 May 2020) |  |  |  |
| --- | --- | --- | --- | --- | --- | --- | --- | --- |
|  | Total | Expected | Excess | Excess as % of expected | Total | Expected | Excess | Excess as % of expected |
| Cancer | 6741 | 6577 | 164 | 2.5 (2.1 to 2.9) | 2552 | 2425 | 127 | 5.2 (4.4 to 6.1) |
| Circulatory | 6541 | 6557 | -16 | -0.2 (-0.1 to -0.4) | 2516 | 2287 | 229 | 10.0 (8.8 to 11.2) |
| Dementia | 3151 | 2735 | 416 | 15.2 (13.9 to 16.6) | 1337 | 897 | 440 | 49.1 (45.8 to 52.3) |
| Respiratory | 2769 | 3462 | -693 | -20.0 (-18.7 to -21.4) | 901 | 1037 | -136 | -13.1 (-11.1 to -15.2) |
| Others | 6297 | 5624 | 673 | 12.0 (11.1 to 12.8) | 2430 | 2004 | 426 | 21.3 (19.5 to 23.0) |

†Cause-specific deaths categorised using ICD-10 codes. These include cancer (C00-C97), dementia/Alzheimer's (F01, F03, and G30), circulatory diseases including heart disease and stroke (I00-I99), non-COVID respiratory diseases (J00-J99), and others.

Because the number of overall deaths was lower than the 5-year average in the period before COVID-related deaths increased in April, we also report excess deaths over the 8-week period when the excess mortality peaked. Between 23^rd^ March and 17^th^ May 2020, there were 48% (N = 9,734) excess deaths, and 13% (N = 1,087) excess non-COVID deaths. Dementia and other causes remained major contributors to the excess deaths (Table 2). Deaths from cancer and circulatory diseases were 5% and 10% higher than expected, respectively. The deficit of deaths in respiratory diseases was –13%.

Changes in non-COVID cause-specific mortality compared with expected mostly occurred between the start of April and the start of May, returning to expected levels thereafter (Figure 3).

**Figure 3.**
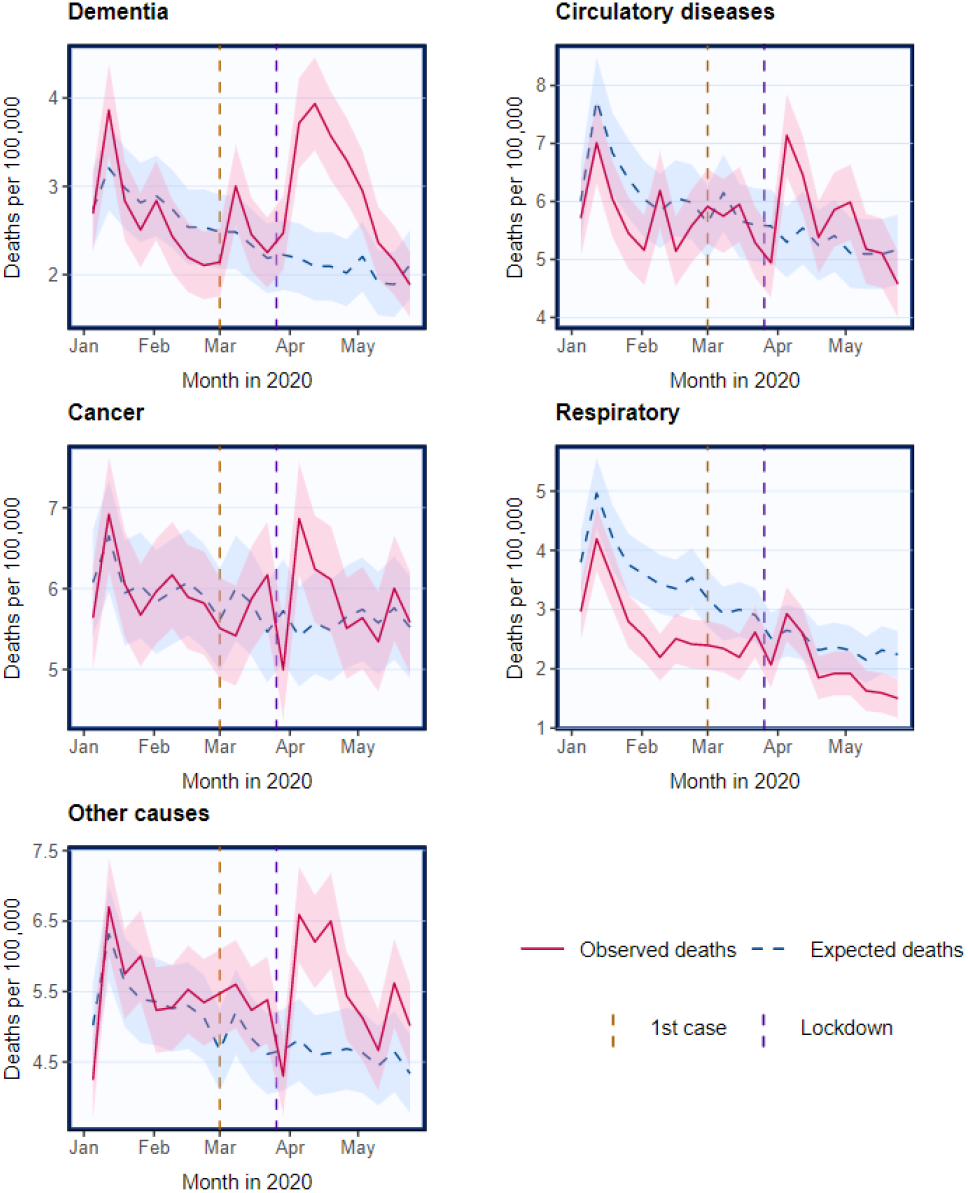
**Crude** mortality trends in Scotland stratified by causes of death

### Publicly available online resource

The mortality trends reported here are changing as the COVID-19 pandemic evolves and both population-wide suppression measures and health services change. Having a responsive and updated portal for visualising and understanding trends should inform policymakers and other stakeholders. We have developed an online tool – TRACKing Excess Deaths (TRACKED) to inform users of the latest mortality trends http://www.trackingexcessdeaths.com). All the findings reported here can be found in the web application, which enables dynamic exploration and visualisation of the data.

## Discussion

COVID-19 was directly responsible for most excess deaths observed in 2020. Using publicly available mortality data in the UK, we also demonstrated an increase in non-COVID excess deaths during the COVID-19 pandemic, which was more pronounced in England and Scotland than in Wales and Northern Ireland. There were excess deaths due to cancer, circulatory disease, dementia, and other causes but not respiratory diseases. Non-COVID deaths have now returned to the expected levels. Overall mortality was lower than expected in early 2020 prior to its steep increase at the beginning of April; interpretation of excess deaths should take this into account. Our online, interactive tool provides the latest trends and figures on excess deaths in all four nations of the UK.

There are several potential explanations for the observed excess of non-COVID deaths. First, demand for SARS-CoV-2 infection testing generally exceeded testing capacity before and during the peak of COVID-19 pandemic. Common symptoms of COVID-19 are also non-specific.6 Hence, even though COVID-19-attributed deaths include suspected as well as confirmed COVID-19 cases, under-diagnosis of COVID-19 (and hence its recognition as a cause of death) is likely and its extent will vary. The observation that the rise and fall of non-COVID deaths closely mirrors that of COVID-related deaths suggests that misclassification of cause of death is likely to contribute to the trends observed.

Second, the definition of COVID-related deaths includes those when COVID-19 was mentioned on the death certificate, whether it was the primary cause, contributing cause, or suspected.2,7 This contrasts with the convention of using the underlying cause of death to categorise cause-specific deaths in population statistics. The definition of COVID-related deaths may mask trends in other causes of deaths, therefore underestimating the impact of COVID-19 on non COVID-related deaths and cause-specific deaths.

Third, many health authorities and clinicians have raised concerns regarding the potential adverse effects of change in health behaviours of the public. Both Public Health England and Scotland have reported a marked reduction in emergency care attendances.8–10 Data from other parts of the world are consistent with the general health behaviours observed in the UK. For example, in the United States, revascularisation for ST-elevation myocardial infarction (STEMI) across nine centres reduced by 38%,11 while time from symptom onset to first medical contact in a single centre in Hong Kong increased from 83 minutes before to 318 minutes during the COVID-19 pandemic.12 It is likely that some patients in the UK have died during this pandemic as a result of not receiving time-sensitive life-saving medical intervention or as a result of delayed presentation to medical services. The return of non-COVID deaths to the expected levels may indicate a change in health behaviour following a public information campaign to persuade the public to seek urgent medical help when required.13

Fourth, some health services had to be suspended to increase capacity for treating people with COVID-19. This involved most non-urgent care, including screening programmes and non-urgent surgery for cancer and other conditions. These restrictions are unlikely to contribute to short-term mortality, which reflects changes in urgent care, but they may have significant impact on medium to long-term mortality, investigation of which will require longitudinal analysis.

As discussed, non-COVID mortality is likely to be under-reported because of the definition of COVID-related deaths. The WHO recommends classifying a death due to COVID-19 as “a death resulting from a clinically compatible illness, in a probable or confirmed COVID-19 case, unless there is a clear alternative cause of death that cannot be related to COVID disease”.14 Reporting of deaths attributable to COVID-19 according to the WHO recommendation as well as other underlying causes will clarify trends of cause-specific excess deaths. More accurate and consistent recording of mortality data during the COVID-19 pandemic will require standardisation of death certification through specific advice for medical practitioners.

### Limitations

First, variable delays in registration of deaths are likely to affect these data. For example, 17% and 27% of deaths were registered more than 7 days after date of death during 2018 in Wales and England, respectively.15 Completeness increased to 92% by 1 month. The current data release lag of 2 weeks by the ONS allows better capture of deaths, though incompleteness is still likely to be about 10%. In Scotland, death registration is more timely (95% within 8 days)16 because of legislation that requires it.17 Whether the COVID-19 pandemic has affected registration delay is currently unknown. Hence, deaths are under-reported and the mortality data used in our analyses provisional, but registries will continue to update their data. Our online tool uses the latest data, incorporating revisions as well as the latest available mortality data.

Second, the current data release does not include age-standardisation or stratification for cause-specific deaths. Should these become available, we will enhance our online tool to provide further information on these. Third, the NRS may under-report mortality data because it does not include deaths occurring outside Scotland. However, the effect of this will be minimised by the full suppression lockdown strategy as most people stay at their usual residence. Fourth, the causes of deaths provided by the NRS are broad and do not allow the identification of high-risk groups most susceptible to the indirect impact of COVID-19. For example, COVID-19 is likely to have a larger impact on more advanced stage and grade cancers.

### Future developments

As data availability increases, we plan to incorporate age- and gender-adjusted mortality data into our online tool to better monitor non-COVID mortality.

Mortality trends stratified by age and gender will help identify those at highest risk. If location-based and cause-specific data become available, we will include these to examine geographical variations and changes in cause-specific mortality.

### Conclusion

There is excess mortality directly and indirectly associated with COVID-19 in the UK. The number of non-COVID excess deaths peaked at the same time as COVID-related deaths but now appear to have returned to the expected levels. Deaths attributed to major non-COVID causes, including dementia, cancer, and circulatory diseases, increased over the peak of the pandemic. Continuous monitoring of these trends and further integration of information on age, gender, location, and cause of death into our online tool will enable increasingly detailed and dynamic assessment of the impact on mortality of the COVID-19 pandemic and associated health policies. The strategy that we have described and observations we have made will be relevant to other countries.

## Data Availability

Links to source codes available in the manuscript. All data used are publicly available.

